# Spatial Cluster Modeling of Rodent Infestations and Leptospirosis Risks in SE Asian Urban Areas

**DOI:** 10.1101/2022.03.24.22272865

**Authors:** Andri Wibowo, Erze Vazela, Alifa A. Prasetyo, Alfian F. Firdaus, Lulu Fajrina, Feby Annisa, Izzah Dinilah, Adinda Nisrina, Fayza Chairunnisa, Farras Putri Aulia, Diah Megakusuma, M. Adrian Dharmawan

## Abstract

Urban areas in the Southeast Asia Region are characterized by rainfall, river networks, and rodent infestations. Combinations of these adverse conditions will lead to the increasing risk of leptospirosis as usually contained in rodents. Then this study aims to assess the spatial pattern of rodent infestations and estimate the potential leptospirosis risks using environmental variables including distance to the river and rainfall in a city in SE Asia. The spatial modeling of rodent infestations was developed based on GIS and interpolation analysis. Meanwhile, the cluster modeling of rodent infestations was developed using the K-means clustering method. The results revealed the rodent infestations represented by two rodent species were *Rattus rattus* and *Rattus norvegicus. R. rattus* has a higher abundance than *R. norvegicus*. In contrast, *R. norvegicus* has wider distribution areas than *R. rattus*. Regarding the distribution areas, both species overlapped in the Southern parts of the city. *R. rattus* and *R. norvegicus* showed a distinct cluster characterized by a high rodent population with affinity for the nearest river, and this indicates the urban inhabitants near the river have more leptospirosis risk. The model of leptospirosis risks estimated an urban area of 35.182 km^2^ or 17.56% having leptospirosis potential.

## 1. Introduction

Rodents, particularly in urban areas, play an important role in Leptospira epidemiology and transmission. Rodents are known to harbor a variety of pathogenic *Leptospira* spp. serovars capable of causing sickness in humans and animals. Because they are numerous in urban areas, rodents commonly found in urban areas include *Rattus norvegicus* and *Rattus rattus* species, which are the most important sources and common reservoirs of Leptospira infection. The prevalence of Leptospira in urban rodents varied greatly depending on geographic location, with some studies reporting zero prevalence in countries including Madagascar, Tanzania, and the Faroe Islands, and others reporting more than 80% prevalence in Brazil, India, and the Philippines (Boey et al. 2019).

Rodent species in urban areas are known to contain Leptospira infections. The 191 strains isolated from urban rodent species belonged to six serogroups including *Leptospira icterohaemorrhagiae, L. bataviae, L. semeranga, L andamana* and *L. ballum*. Determinant factors influencing the Leptospira infection among urban rodents include the age of the rodents and the area in which they are found (Lindenbaum &Eylan, 1982).

Rodent associations and the prevalence of Leptospira are also on the horizon in SE Asia Regions. In these regions, leptospirosis is thought to be endemic in Vietnam, Cambodia, and Laos. Research conducted in 2002 by Laras et al. proved the disease’s endemicity in several locations in those countries. In this investigation, Hurstbridge, Bataviae, and Icterohaemorrhagiae were found to be the most common serovars in patients with clinical jaundice. In the Philippines, poor sanitation, an increase in urban slums, as well as frequent typhoons and the extension of flood areas in the country, have all contributed to an increase in the risk of infection (Liongson et al. 2000). Flooding was a major cause of leptospirosis outbreaks in prisons, penal farms, and other areas throughout the Philippines (Basaca-Sevilla et al. 1986).

Indonesia is also one of the countries in the SE Asia Region that is threatened by Leptospira epidemiology and transmission in urban areas. Rodents infected by Leptospira were recorded in a wide array of urban areas. A rodent sample collected from various locations in urban areas, ranging from a settlement to a harbor, confirmed a rodent individual containing Leptospira. Leptospirosis is most likely a serious and underappreciated ongoing health problem in Indonesia. In 2001, among 139 human serum samples, 18.7% of the samples tested positive for containing Leptospira. Since 2006, there has been a countrywide upsurge in the number of reported human cases. In 2007, 93% of 667 reported human cases were laboratory confirmed.

The presence of leptospirosis in the environment is quite difficult to detect, considering Leptospira is a microscopically organism and hard to detect. Despite these obstacles, because the Leptospira carrier is a rodent, detecting the presence of rodents will provide insight into the risk of leptospirosis disease potential. GIS-based approach and spatial analysis have been developed and implemented to model the spatial distributions of rodent individuals along with the estimations of leptospirosis disease potential. A GIS-based approach has been used to anticipate the disease spread related to *R. norvegicus, R. rattus*, and human encounters. Based on spatial analysis, it is confirmed that urban slums (84.5%) are at the highest risk, followed by croplands (10.9%) and shrublands (2.7%). Anthropogenic factors affecting the incidence of rodent encounters included human density, distance from water channels, land use/land cover, and distance from roads (Fatima et al. 2018).

### 1.1. Research significance

Depok, like other tropical SE Asian cities, is receiving heavy rainfall, especially during La Nina. Previous research has reported rodent infestations and emergences of leptospirosis cases after the rainy season. Meanwhile, the information about the rodent infestations and potential for Leptospira epidemiology and transmission in Depok is still limited. Lack of data on rodent infestations hinders the delineation of urban areas in Depok City that might be threatened by Leptospira epidemiology and transmission. Then this study aims to map the spatial distributions of rodent infestations and estimate the potential areas might be threatened by leptospirosis.

## 2. Methods

Methodology for this study is following Lau et al. (2012) and Radi et al. (2016).

### 2.1. Study area

The study area was located in Depok, a growing satellite city at a distance of 50 km from the capital City of Jakarta (Figure 1). The size of Depok was 200.3 km^2^ and had a total population of 2,719,813 people with density of 5,982 people/km^2^. The topography of Depok is ranging from 50 to 140 m asl. Depok is receiving a high rainfall with averages of 327 mm and the highest is 591 mm.

**Figure 1.**
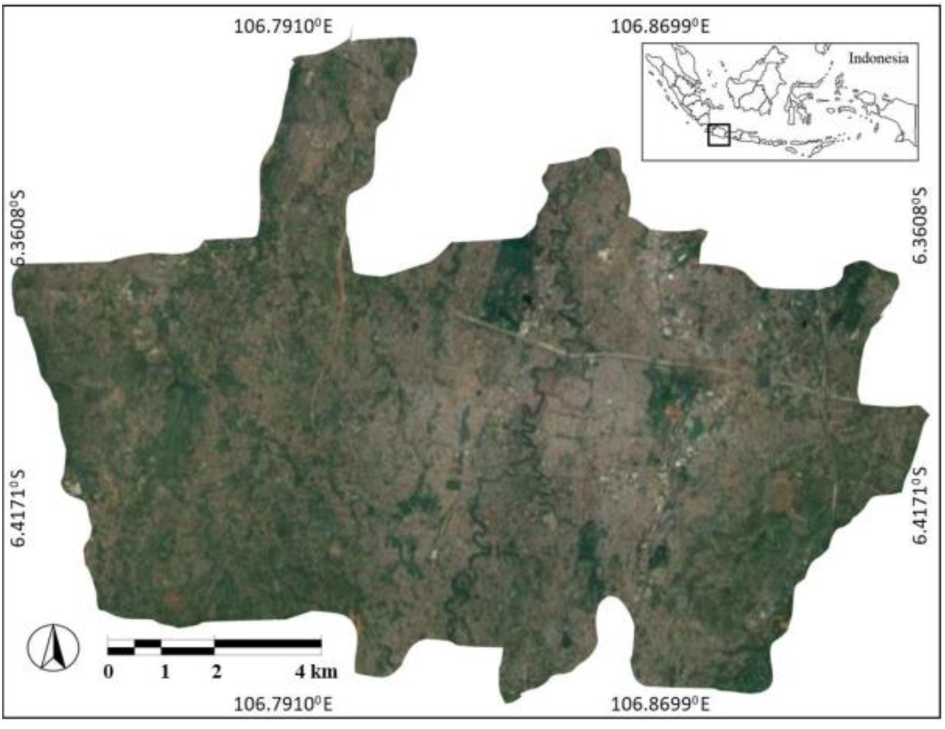
Study area (Google Map, Terra Metric).

### 2.2. Rodent observations

The rodent observation in the urban areas in this method was based on visual and audio encounters. Visual observation or sighting is applied to rodent species that inhabit terrestrial ecosystems. Meanwhile, audio encounter was applied to rodent species that inhabit above-ground ecosystems, including roofs.

### 2.3. Rodent study areas and environmental variable mapping

Rodent, study area, and environmental variable mappings were based on GIS methodology. First, the locations of rodent encounters were recorded using GPS to determine their latitude and longitude. The rodent data was then tabulated in a geoattribute table and represented as points in the GIS environments. The point data of rodents is then interpolated to estimate the spatial distribution of rodents.

The satellite image of the study area was obtained and classified into environmental variables related to the rodent population. Those environmental variables include rivers and rainfall. The river was represented as a polyline and the rainfall as a polygon. For analysis purposes, the spatial data of interpolated rodent populations was overlaid with river and rainfall data using GIS. The study area (Figure 1) was used as a background image.

### 2.4. Rodent population cluster model

Cluster methods aim to determine the concentrations and hotspots of the rodent population in studied city follow current methods (Grubesic &Murray, 2001). The input data were the abundances of rodent population and sighting in each sampling point and were presented as points in the GIS interface. Cluster analysis was conducted using an extension of GIS and the cluster calculation was based on the K-means method. This method uses an algorithm that assigns each point to the cluster whose center or known centroid is nearest (Aksoy, 2006). The center is the average of all the points in the cluster, and the coordinates of the points are the arithmetic mean for each dimension separately over all the points in the cluster. The determination of the centroid, or cluster point, was as follows:

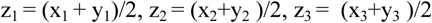

with: z = centroid, x = coordinate in axis x, y = coordinate in axis y

### 2.5. Leptospirosis spatial model

A leptospirosis spatial model was developed based on the ranking and overlay of environmental variables that contributed to the leptospirosis risk potential. All the environmental variables and rodent populations for observed variables were ranked from the lowest to the highest. The lowest rank of variables was considered low-risk, and the highest rank of variables was considered high-risk. Then all the rank values were tabulated into the GIS table along with their geocoordinates. Furthermore, the tabulated GIS tables contained environmental variables and rodent population variable ranks were used and interpolated to create GIS layers and vector shapes. All the layers were overlaid and the rank values in each variable layer and vector shape were summed up to create composite layers. The final step was to classify rank values in composite layers, and the vector shapes with the highest rank values were classified as the most leptospirosis risk areas. While the vector shapes with the lowest rank values were classified as having the least leptospirosis risk.

## 3. Results and Discussion

### 3.1. Rodent infestations

The observations and sightings data on rodents in this study confirmed 2 rodent species, they were *Rattus rattus* and *Rattus norvegicus*. The presences of both rodent species have similarity since both species are observed in the Southern parts of Depok. The differences were the coverage and spatial distributions of those species. *R. norvegicus* species were observed have wider coverage and spatial distributions in the Southern parts of Depok (Figure 2). In contrast, *R. rattus* (Figure 3) have smaller coverage and spatial distributions than *R. norvegicus*. The *R. rattus* is a commensal species that inhabits environment thrives alongside humans settlement throughout the world. *R. rattus* species being less often associated with human activity than *R. norvegicus* that were found in wider urban areas (Harper &Bunbury, 2015). In contrast, *R. norvegicus* was less abundant than *R. rattus* and virtually absent in natural environments where human pressure is low. Our sighting data confirm despite more *R. rattus* individuals were observed in urban areas, this species has limited coverage. This is consistent with the fact that *R. norvegicus* species were the superior rodent competitors in urban habitats and limit the coverage of *R. rattus*.

**Figure 2.**
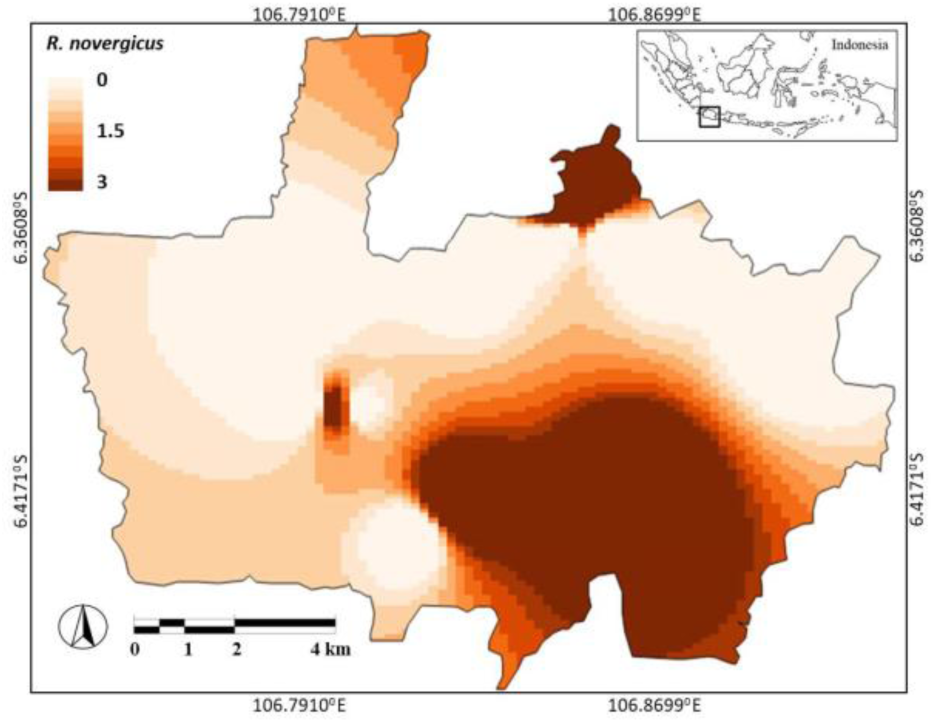
Interpolated and estimated *Rattus norvegicus* sightings in urban areas.

**Figure 3.**
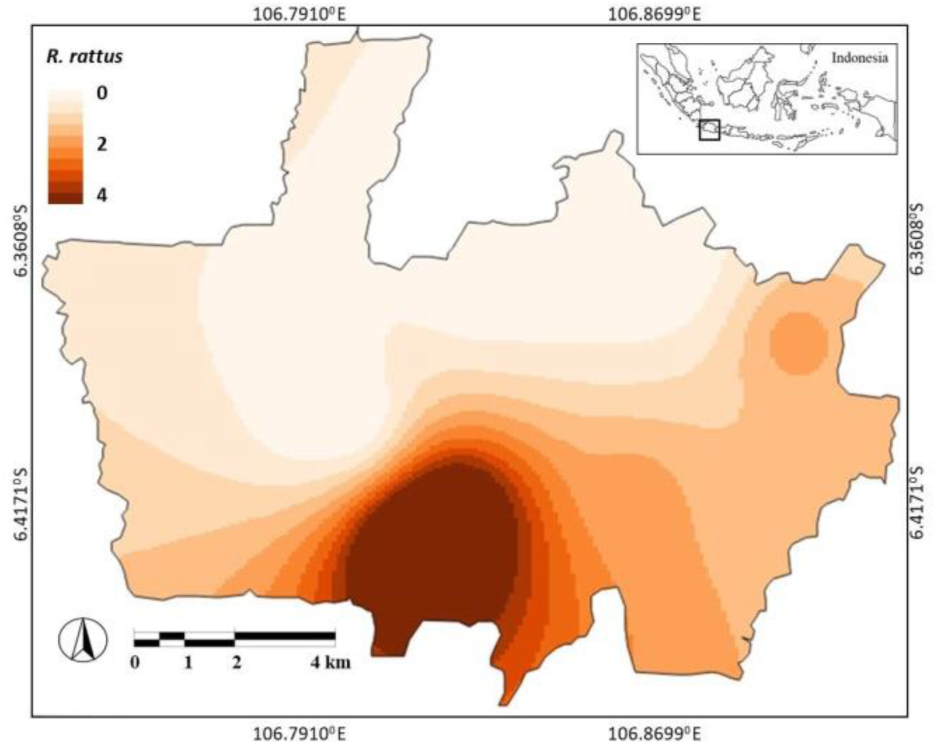
Interpolated and estimated *Rattus rattus* sightings in urban areas.

Wider coverage and spatial distributions of *R. norvegicus* then indeed raise and health issue regarding the leptospirosis disease transmission. Previous research confirmed that *R. norvegicus* species were observed have higher leptospira strains than *R. rattus*. Leptospires were isolated from 21.2% of *R. norvegicus*, meanwhile only 9.9% of *R. rattus* has leptospira isolates. Even in contrast, *R. tanezumi*, a species closely related to *R. rattus* and a shrew species were infected more by Leptospira (Widiastuti et al. 2016).

### 3.2. River network and rainfall pattern in urban areas

The study areas were passed by 2 rivers in the East and West in a South-North direction. The river in the East was larger than in the West (Figure 4). The study area has 3 types of rainfall. The rainfall pattern was increasing toward the South. Low rainfall was observed in the Northern parts of the city. In contrast, the Southern parts of the city are receiving the highest rainfall, amounting to almost 350 mm.

**Figure 4.**
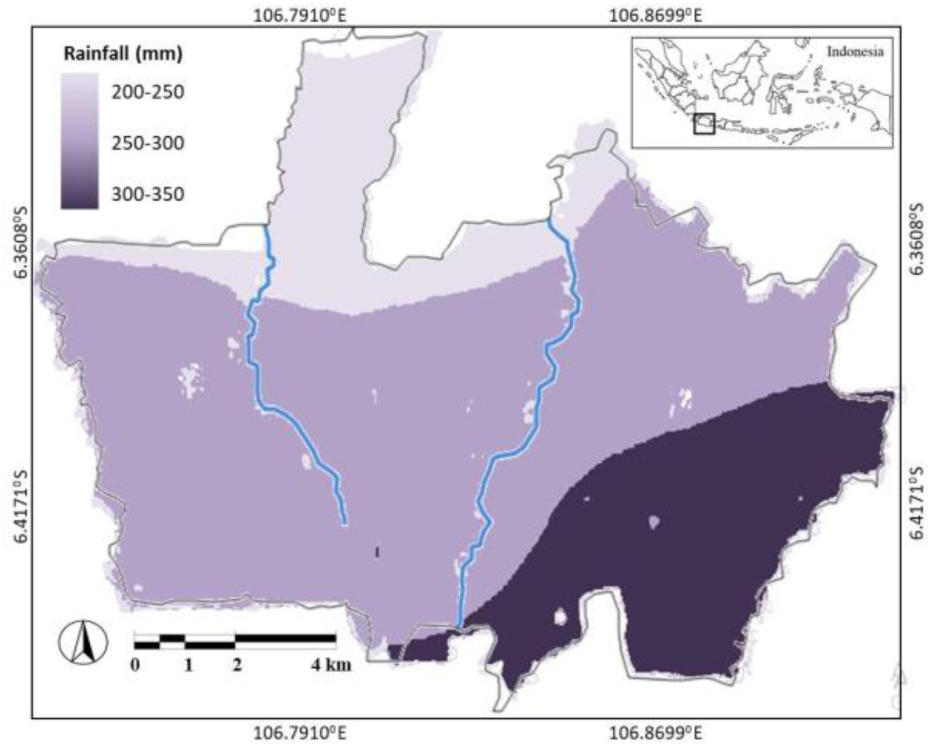
River network and rainfall pattern in urban areas.

### 3.3. Rainfalls and rodent populations

The spatial model shows the Southeast parts of Depok receiving the highest rainfall in comparison to other parts. Rainfall patterns were observed to increase in the South, with the Northern parts of the city receiving the least rainfall. This pattern coincided with the patterns of rodent infestations. *R. norvegicus* was found to have high populations in the Southern parts. In comparison, the population of *R. rattus* species also increased in the Southern parts. Rats prefer wet areas indicated by high rainfall. First, wet conditions are important to support the reproduction period for rats. Wet conditions due to rain will increase soil moisture and decrease the heat stress of rats. Rats are in excellent physical and vigorous condition to reproduce and hence can breed over an extended period, which as a result leads to an increase in rodent populations (Madsen & Shine, 2009). In this suitable microhabitat, rats can reproduce as much as 5-7 times a year.

The correlation of *R. rattus* with the coverage of areas receiving high rainfall in this study is in agreement with results from other studies. Rainfall actually has an indirect association with rodent populations. If an area has poor drainage, then rainfall will be one of the potential factors contributing to flooding. In fact, flooding, as described in many studies, is a precursor of rodent population increases in urban areas. First, floods will wash away food remnants, including waste, and this can become a potential food resource for rodents (Victoriano et al. 2009). In 2008 in Bhutan, a surge in rodent populations was observed at the onset of summer after flooding. In Indonesia, the wake of massive flooding in Indonesia in January 2002 was followed by a surge in rodent populations and a leptospirosis outbreak. As a result of this outbreak, 12.0% out of 418 samples were seropositive against serovars Bataviae or Hardjo.

### 3.4. River streams and rodent populations

The cluster analysis using K-means showed that the *R. rattus* population is divided into 3 clusters (Figure 5). The centroid of the cluster confirmed that the *R. rattus* with a high population were clustered near the river. Meanwhile, for *R. norvegicus*, there were 4 clusters. In comparison to *R. rattus*, the high population of *R. norvegicus* was also clustered near the river.

**Figure 5.**
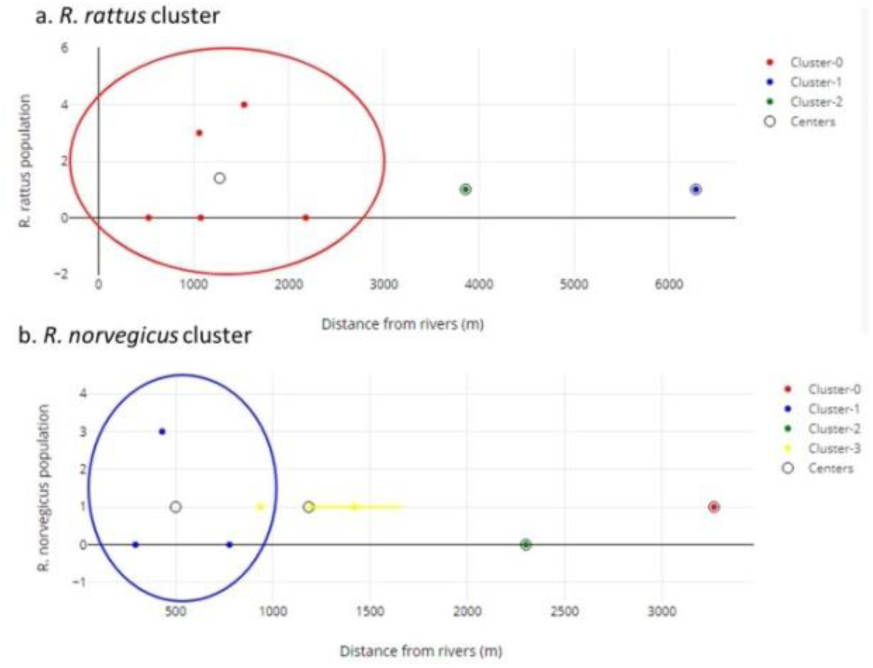
K-means clustering and centroid positions of a. *R. rattus* (top) and b. *R. norvegicus* (bottom) related to distance to river variables.

The associations of rodent presence with the river are related to a number of factors. First, river provides a food resource for the rodents, whose food is washed away from settlements during rain and floods. In urban areas, sometimes there is a slum near the river. This slum provides food for nearby rodent populations and increases the prevalence of leptospirosis cases (Costa et al. 2014; Hagan et al. 2016).

### 3.5. Leptospirosis risk model

Figure 6 estimates the leptospirosis risk model. The model classifies the study area into high, medium, and low risks. From the model, it can be seen that the areas that were estimated to have high risks are located in the Southern parts of the city. The North, West, East, and Central parts of the country had low to medium risks of leptospirosis. Based on the calculations, it is estimated that the size of areas having high risks was 35.192 km, or equal to 17.56% of the city.

**Figure 6.**
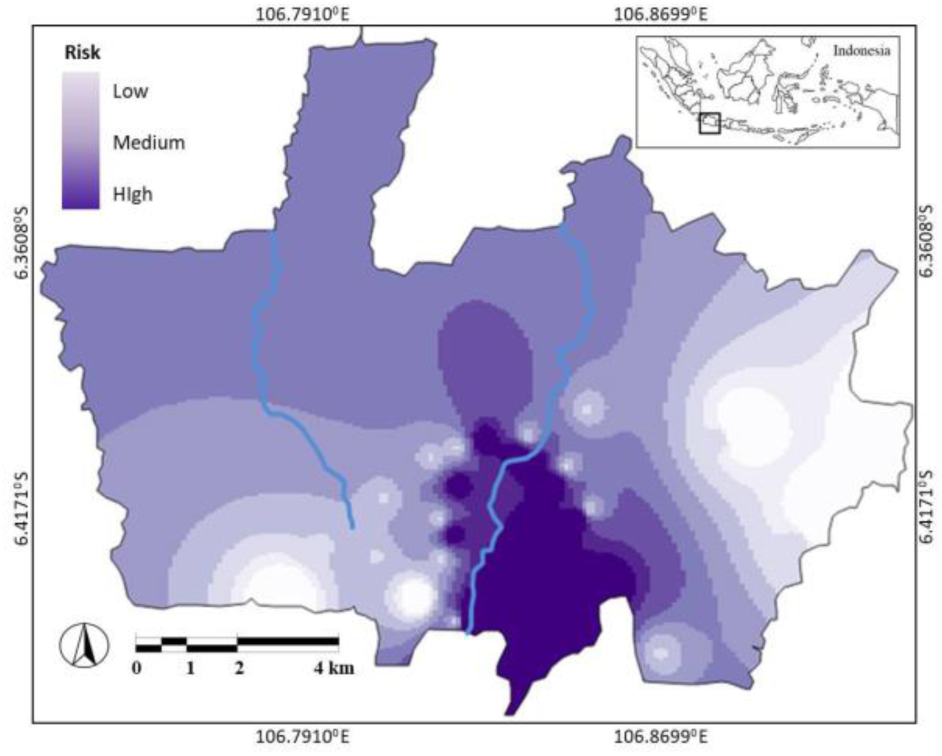
Leptospirosis risk model.

The model shows that areas with high leptospirosis cases are in affinity to the river. This is in consistent with the previous study (Fajriyah et al. 2017) on the associations of the diseases with the presence and distance to the river nearby. In another city in Indonesia, the related risk factors were contact with stagnant water, contact with river or flood water, muddy areas, and also having activities in water including swimming, bathing, washing. Those activities and contact with water can increase contact with animal urine, bodies, or tissues due to not wearing personal protection equipment, walking barefoot, and using streams as a source of drinking water (Sakundarno et al. 2014).

Regarding rainfall variables, the model shows the area with leptospirosis risk is located near area that has high rainfall in Depok City. This is in an agreement with previous research showing associations of rainfall with leptospirosis cases. The association of leptospirosis with rain is mentioned in several publications and measured. In a few others, even large outbreaks have been reported during seasonal periods of heavy rainfall and flooding (Barcellos &Sabroza, 2001; Sarkar et al. 2002). In Thailand, it is observed that outbreaks of leptospirosis correspond with the rainy season, with an increase in cases beginning in August and decreasing in November.

## 4. Conclusions

Leptospirosis remains a major endemic environmental disease in most cities and urban areas in the Southeast Asia region. Distance to the river and rainfall were observed as the potential determinants of the incidence of the rodents that lead to the prevalence of the disease. This study confirmed the rodent infestations in urban areas that have proximity to the river and areas receiving rainfall.

The rodent infestations indicate three potential risks of leptospirosis. The areas considered at risk have a size of 35.192 km^2^, or equal to 17.56% of the city. This area is identified as located in the Southern parts of the city. The leptospirosis risk pattern is increasing southward. Then the results of this study contribute more to the urban and health planners’ ability to monitor and mitigate leptospirosis in a more effective manner.

## Data Availability

All data produced in the present work are contained in the manuscript

## Notes

### Competing Interest Statement

The authors have declared no competing interest.

### Funding Statement

This study did not receive any funding

## References

Aksoy E. Clustering with GIS: an attempt to classify Turkish District data. 2006.

Barcellos C, Sabroza PC. 2001. The place behind the case: leptospirosis risks and associated environmental conditions in a flood-related outbreak in Rio de Janeiro. C 17: 59–67

Basaca-Sevilla V, Cross JH, Pastrana E. 1986. Leptospirosis in the Philippines. Southeast J Trop Med Pub Hlth. 17:71–74

Boey K, Shiokawa K, Rajeev S. 2019. Leptospira infection in rats: A literature review of global prevalence and distribution. PLoS Negl Trop Dis. 9;13(8):e0007499. doi: 10.1371/journal.pntd.0007499. PMID: 31398190; PMCID: PMC6688788.

Costa F, Ribeiro G, Felzemburgh R, Santos N, Reis R. 2014. Influence of Household Rat Infestation on Leptospira Transmission in the Urban Slum Environment. PLoS neglected tropical diseases. 8. e3338. 10.1371/journal.pntd.0003338.

Fajriyah S, Udiyono A, Saraswati L. 2017. Environmental and Risk Factors of Leptospirosis: A Spatial Analysis in Semarang City. IOP Conference Series: Earth and Environmental Science. 55. 012013 10.1088/1755-1315/55/1/012013.

Fatima S, Zaidi F, Adnan M, Ali A, Jamal Q, Khisroon M. 2018. Rat-bites of an epidemic proportion in Peshawar vale; a GIS based approach in risk assessment. Environmental Monitoring and Assessment. 190. 10.1007/s10661-018-6605-7.

Grubesic T, Murray A. Detecting hot spots using cluster analysis and GIS. 2001.

Harper GA, Bunbury N. 2015. Invasive rats on tropical islands: Their population biology and impacts on native species. Global Ecology and Conservation, 3, 607–627.

Hagan J, Moraga P, Costa F, Capian N. 2016. Spatiotemporal Determinants of Urban Leptospirosis Transmission: Four-Year Prospective Cohort Study of Slum Residents in Brazil. PLoS neglected tropical diseases. 10. e0004275. 10.1371/journal.pntd.0004275.

Laras K, Cao BV, Bounlu K, Nguyen TK, Olson JG Thongchanh S, Tran NV, Hoang KL, Punjabi N, Ha BK, Ungsa SA, Insisiengmay S, Watts DM, Beecham HJ, Corwin AL. 2002. The importance of leptospirosis in Southeast Asia. Am J Trop Med Hyg. 67:278–286.

Lau C, Clements A, Skelly C, Dobson A. 2012. Leptospirosis in American Samoa – Estimating and Mapping Risk Using Environmental Data. PLoS neglected tropical diseases. 6. e1669. 10.1371/journal.pntd.0001669.

Liongson LQ, Tobias GQ, III, Castro PPM. 2000. Pressure of urbanization: flood control and drainage in Metro Manila. University of the Philippines Center for Integrative Developments Studies;.

Lindenbaum I, Eylan E. 1982. Leptospirosis in Rattus norvegicus and Rattus rattus in Israel. Isr J Med Sci. 18(2):271–5. PMID: 7068359

Madsen T. Shine R. 2009. Rainfall and rats: Climatically-driven dynamics of a tropical rodent population. Australian Journal of Ecology. 24. 80 –89. 10.1046/j.1442-9993.1999.00948.x.

Radi M, Jaafar M, Hod R, Ahmad N. 2016. Leptospirosis outbreak following the 2014 major flooding event in Kelantan, Malaysia-a spatial-temporal analysis.. ISEE Conference Abstracts. 2016. 10.1289/isee.2016.4580.

Sakundarno M, Bertolatti D, Maycock B, Spickett J, Dhaliwal S. 2014 Risk factors for leptospirosis infection in humans and implications for public health intervention in Indonesia and the Asia-Pacific region. Asia. Pac. J. Public Health 26 15–32.

Sarkar U, Nascimento SF, Barbosa R, Martins R, Nuevo H, Kalofonos I, Kalafanos I, Grunstein I, Flannery B, Dias J, Riley LW, Reis MG, Ko AI. 2002. Population-based case-control investigation of risk factors for leptospirosis during an urban epidemic. Am. J. Trop. Med. Hyg. 66 605–10.

Victoriano, AF. Smythe LD, Gloriani-Barzaga N, Cavinta L L, Kasai T, Limpakarnjanarat K, Ong BL, Gongal G, Hall J, Coulombe CA, Yanagihara Y, Yoshida S, Adler B. 2009. Leptospirosis in the Asia Pacific region. BMC infectious diseases, 9, 147. https://doi.org/10.1186/1471-2334-9-147.

Widiastuti D, Sholichah Z, Agustiningsih A, Wijayanti N. 2016. Identification of pathogenic leptospira in rat and shrew populations using RpoB Gene and its spatial distribution in Boyolali District.

